# Hepatitis B virus infection status and associated factors among health care workers in selected hospitals in Kisumu County, Kenya: a cross-sectional study

**DOI:** 10.1101/2023.01.06.23284271

**Authors:** Frankline Otieno Mboya, Ibrahim I. Daud, Raphael Ondondo, Daniel Onguru

## Abstract

**Background:** Poorly managed medical waste produced at the health facilities are potential source of infections including occupational exposure to Hepatitis B Virus (HBV). This study evaluated the prevalence of HBV infection among healthcare workers (HCW) in Kisumu County.

**Methods:** We determined the prevalence of HBV infections among 192 HCW from nine purposively selected high volume public hospitals in Kisumu County. A structured questionnaire was administered, and 4.0 ml of venous blood sample collected for Hepatitis B surface antigen (HBsAg), hepatitis B surface antibody (anti-HBs) and total hepatitis B core antibody (anti-HBc) testing using enzyme immunoassay (EIA).

**Results:** Of the 192 HCW sampled, 52.1% were males and 78.7% are married, the median participants age was 34.4 years with interquartile range (IQR) of 11 years. Most participants had between 1-5 years of service while 43.8% had ≥2 doses of HBV vaccine.

The respective prevalence of HBsAg, anti HBs and anti HBc was 18.8% (95% CI: 13.5-25.0%), 63.0% (95% CI: 55.8-69.9%) and 44.8% (95% CI: 37.6-52.1%). Higher proportion of HBV positive was found in HCW who had worked for less than one year and who had not received any dose of HBV vaccine at 37.5% and 35.9 % respectively.. Significant risk of HBV lifetime exposure was noted among HCW with one vaccine dose, those with no known exposure and highest in those with knowledge on HBV transmission (aOR, 7.97; 95% CI, 2.10-153.3, p-value=0.008). HCW who had received ≥2 doses of HBV vaccine (aOR, 0.03; 95% CI, 0.01-0.10, p-value= <0.0001) had significant HBV protection. Duration of service was not associated with HBV among HCW.

**Conclusion:** Prevalence of HBV infection was high among HCW in Kisumu County. Ministry of health Kenya should strengthen comprehensive infection prevention and control practices to reduce lifetime exposure to HBV infection among HCW.

## Background

Globally, it is estimated that 257 million people are living with chronic hepatitis B (CHB) viral infection, HBV epidemic mostly affects WHO African and Western Pacific Regions (1). Prevalence of HBV infection in Africa is averagely more than 10% while pooled HBV prevalence in HCW was 6.81% (95% CI 5.67–7.95) classifying the region as one of high endemic area (2–5). In 2007, the Prevalence of HBV infection in Kenya was estimated to be 2– 5%, while 31% of the Kenyan population was found to have been previously exposed to HBV (6).

Global mortality due to viral hepatitis was about 1.4 million in 2016 (7, 8), of those deaths approximately 47% were due to HBV infection (9). HBV is highly infectious and is mainly transmitted through vertical transmission, percutaneous blood, sexual and body fluid contacts(10). Health care services in hospitals is aimed at restoring health and saving lives (11), these services also generate infectious medical waste that if poorly managed could be potential source for hospital acquired infections (HAI) which include HBV (12). The risk of HAI increases when basic infection prevention and control (IPC) practices in health care settings are not well laid out and adhered to. There is limited data on occupational exposure to HBV infection and its prevalence among HCW in Kenya. This study estimated the prevalence of HBV infection and exposures among HCW in Kisumu County.

## Methods

### Study aim, design, and setting

We conducted descriptive cross-sectional study between May 2020 and April 2021 to estimate HBV infection prevalence and risk factors among HCW in nine largest public hospitals in Kisumu County. These hospitals provide primary and referral medical care services, trainings, and have different structural establishments depending on the hospital level. The hospitals generate varying nature and type of medical waste depending on their capacity and medical services/procedure offered.

### Study population and characteristic of the participants

A total of 192 HCW (nursing officers, medical officers, clinical officers, medical laboratory technologists, medical waste handlers, HIV testing counsellors, and mortuary technicians) were included in the study. Probability proportional to size (PPS) sampling was used to identify number of participants in each selected health facility and service delivery points, simple random sampling from duty roster was used to sample the participants.

### Data collection and Laboratory Procedures

A structured questionnaires was administered to collect sociodemographic data after obtaining informed consent. 4.0 ml of venous blood sample was collected in ethylene-diamine-tetra-acetic acid (EDTA) (Becton, Dickinson and Company, Franklin Lakes, New Jersey, USA) for evaluating HBV infection based on three biomarkers: hepatitis B surface antigen (HBsAg), antibodies against hepatitis B surface antigen (anti-HBs), and antibodies against total hepatitis B core antigen (anti-HBc). Current HBV infection was determined by testing for HBsAg using Murex HBsAg version 3 kit; immunity to HBV infection was established by testing for anti-HBs using ETI-AB-AUK-3 Diasorin anti-HBs EIA kit; and past exposure to HBV infection was assessed by testing for anti-HBc using Murex anti-HBc (total) kit. All tests were done as per the manufacturers’ kit instructions without modifications. HBV current infection was defined as individual’s blood is serologically positive for HBsAg while HBV lifetime exposure are individuals whose blood is serologically positive for either HBsAg (current infection) or anti-HBc (may indicate a current or past resolved infection).

### Data analysis

Data was analysed using SPSS version 16.0 (SPSS Inc., Chicago, IL, USA). Results were summarized using descriptive statistics. Logistic regression models were used for bivariate and presented as odd ratios (OR) with 95% confidence intervals (CI). Multivariable analyses were performed for factors attaining p-values ≤0.2 in bivariate analysis to determine independent factors associated with HBV infection (positive for HBsAg or anti-HBc) among HCWs and presented as adjusted OR (aOR). A threshold p-value of less than 0.05 was considered statistically significant. The models were adjusted for age and gender.

### Ethical considerations

The study received ethical approval form JOOTRH ethics review board IERC/JOOTRH/244/20 and National Commission for Science, Technology & Innovation (NACOSTI) granted research permit to conduct the study (licence no: NACOSTI/P/20/6300). Permissions to collect data from hospitals within Kisumu County was granted by Kisumu County Director of Health. All HBV susceptible HCW were referred for HBV vaccination through Kenya Expanded program for immunization (KEPI).

## Results

### Sociodemographic characteristic of HCW

Of the 192 HCW sampled, 52.1% were males and 78.7% are married, the median participants age was 34.4 years with interquartile range (IQR) of 11 years. Most participants had between 1-5 years of service. Varied results were observed in HCW IPC training and capacity building: while capacity building on personal protective equipment (PPE) usage was 73.4%, trainings on waste management was 30.2% and infectious agents found on waste was 32.3%.

There was moderate knowledge on HBsAg transmission, prevention/control and waste disposal at 56.8%, 53.7% and 30.2% respectively. HBV vaccine completion rate was low at 43.8% of HCW receiving ≥2 vaccine doses while 40.6% being susceptible to HBV infection. Whereas 90.6% of health care workers agreed that PPEs are generally available within the work settings, 77.6% felt that they were inadequate. There was low daily usage of PPEs: Apron/dust coat 65.1%, gumboots (waste handlers) 13.5%, gloves 44.8% and mask wearing while on duty 56.3%. There was good access to hand hygiene 96.4% and availability of waste management bins (black, yellow, red) 95.8% with low waste segregation at 29.7% while 67.2 % of waste were incinerated at site or networked to facilities with incinerator. Higher proportion (53.1%) of HCW had either contact or needle stick injury exposure (Table 1).

**Table 1:**
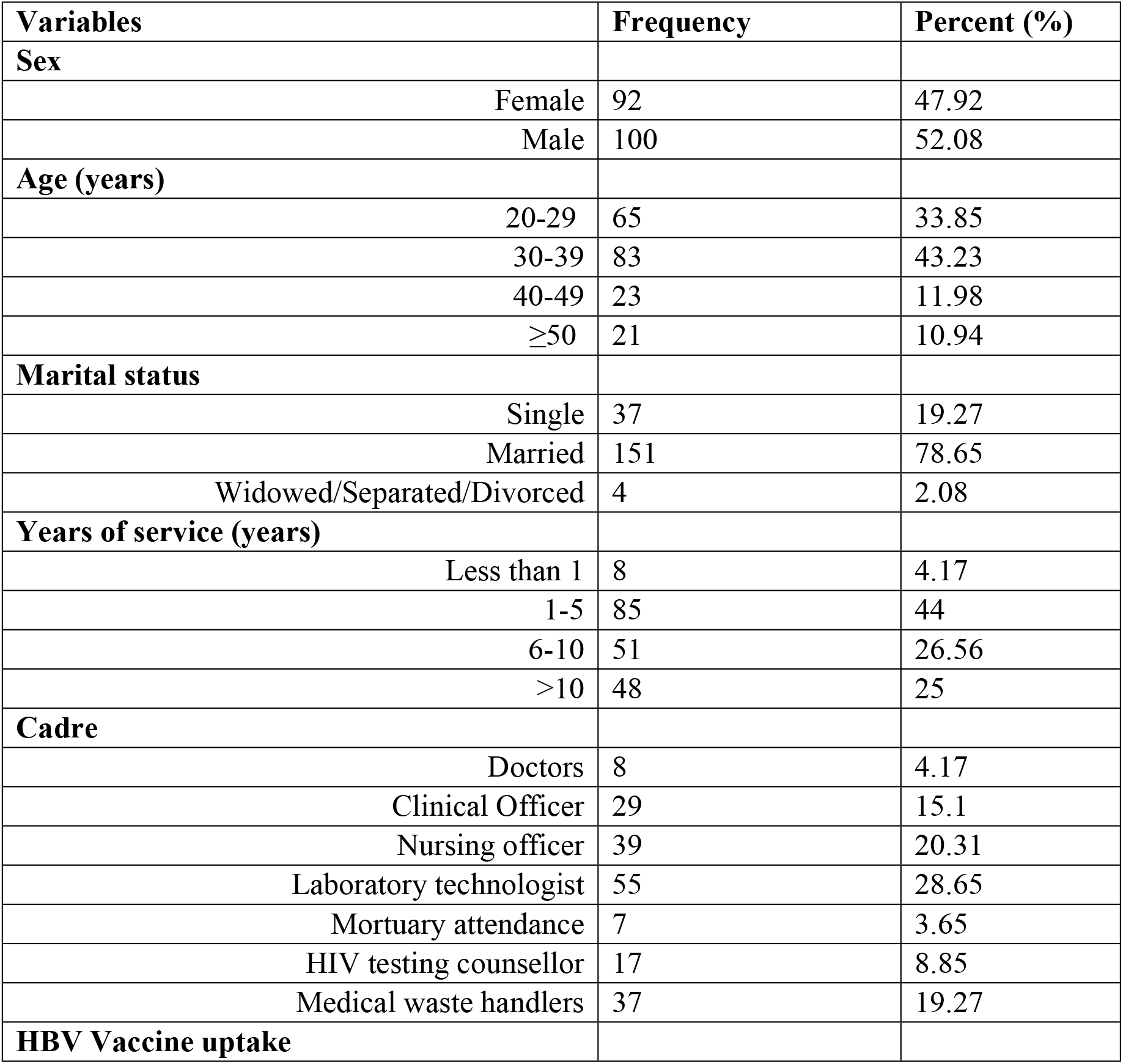
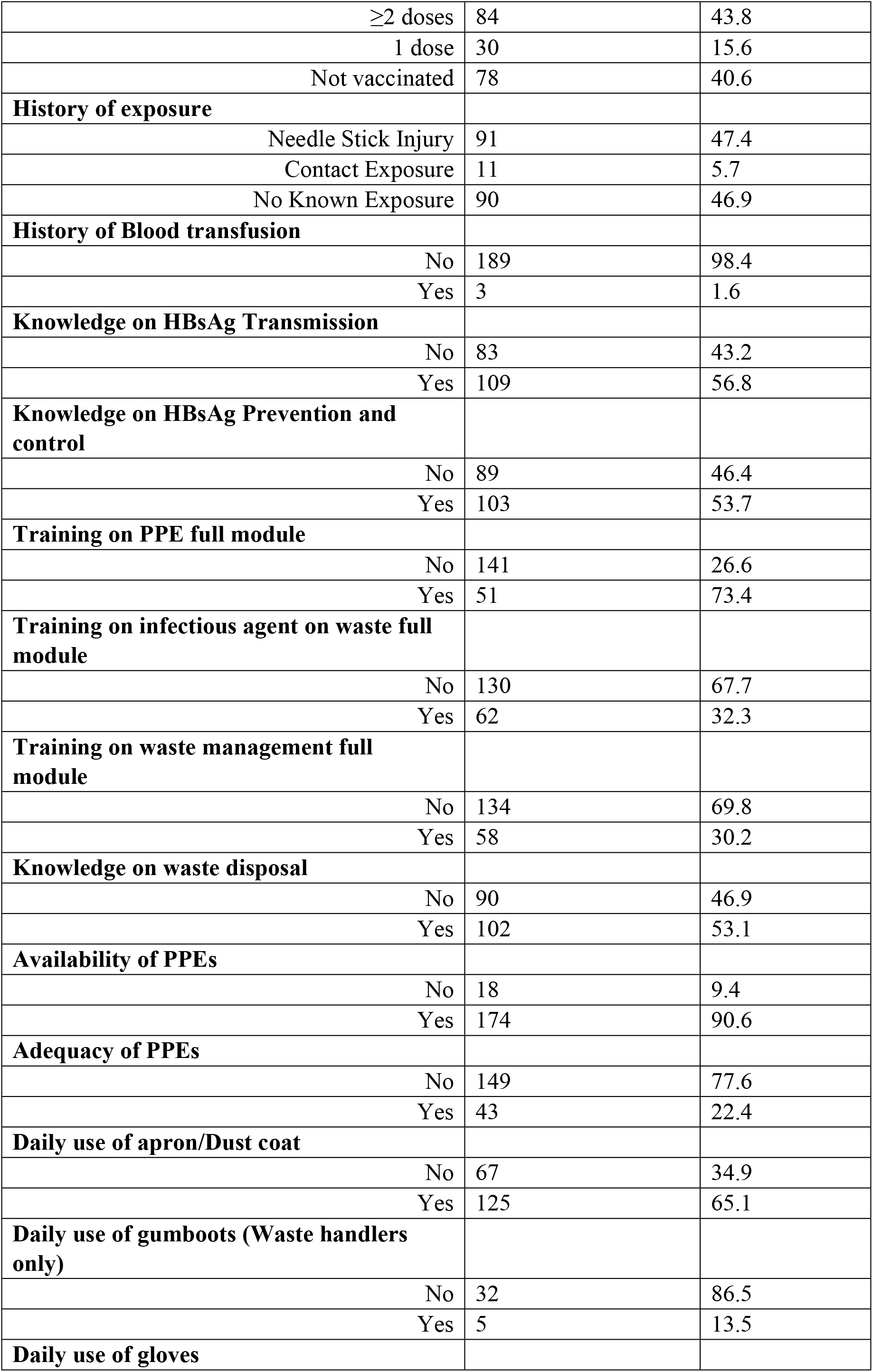
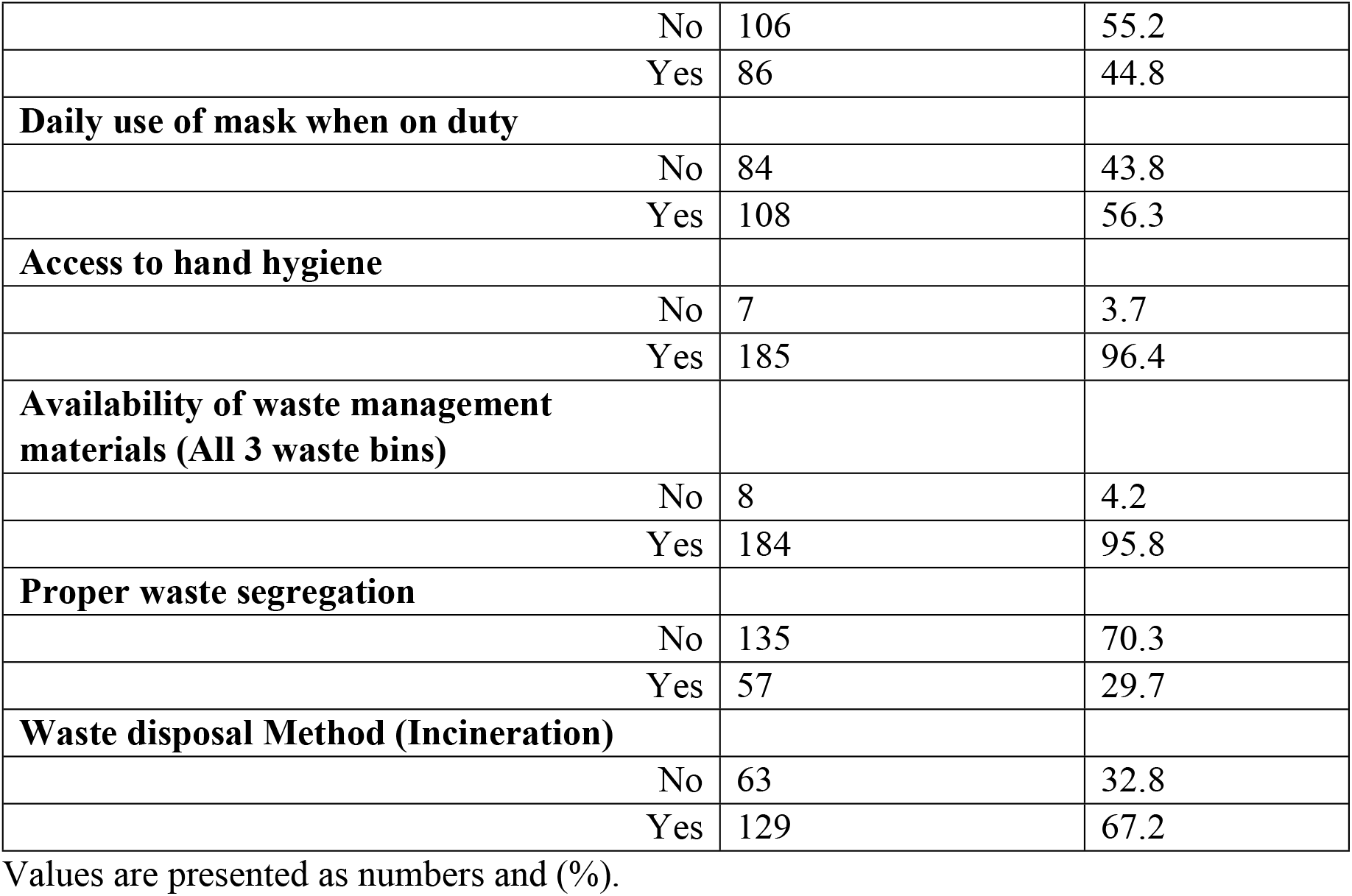
Socio-demographic characteristics and risk factors of the study population (N= 192)

### Prevalence of HBV Biomarkers among HCW in Kisumu County, 2020 (N= 192)

In table 2 below, the respective prevalence of HBsAg, anti HBs and anti HBc was 18.8% (95% CI: 13.5-25.0%), 63.0% (95% CI: 55.8-69.9%) and 44.8% (95% CI: 37.6-52.1%) respectively. Highest prevalence of HBsAg was seen among, HCW who had worked for less than one year 37.5% (95% CI: 8.5-75.5), HBV unvaccinated HCW 35.9% (95% CI: 25.3-47.6), HCW who has had blood transfusion 33.3% (95% CI: 0.84-90.6) and HIV testing counselors 29.4% (95% CI: 10.3-56).

**Table 2:**
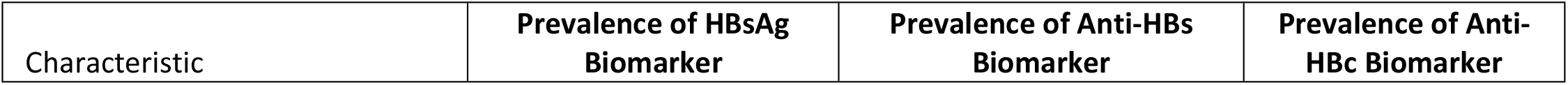
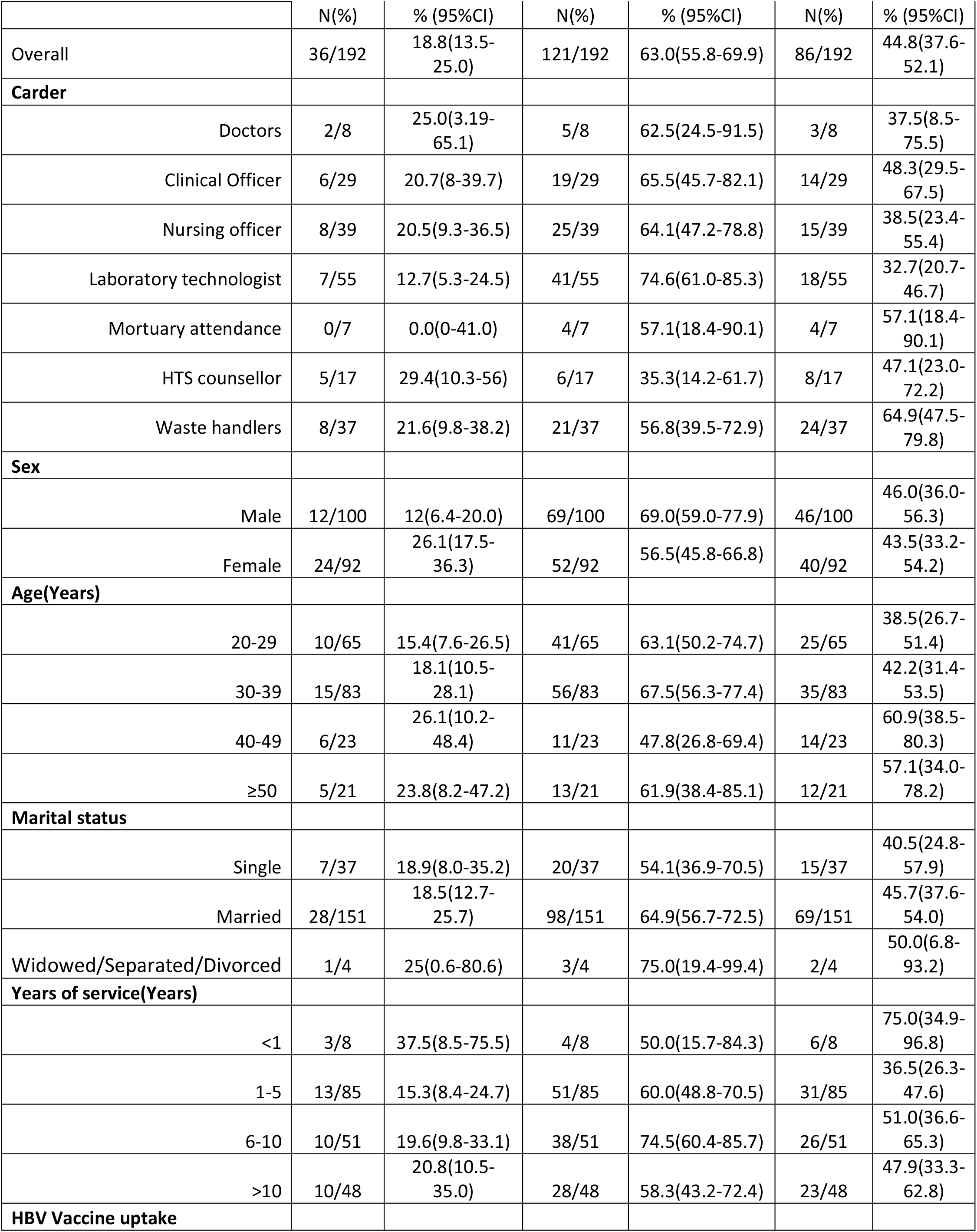
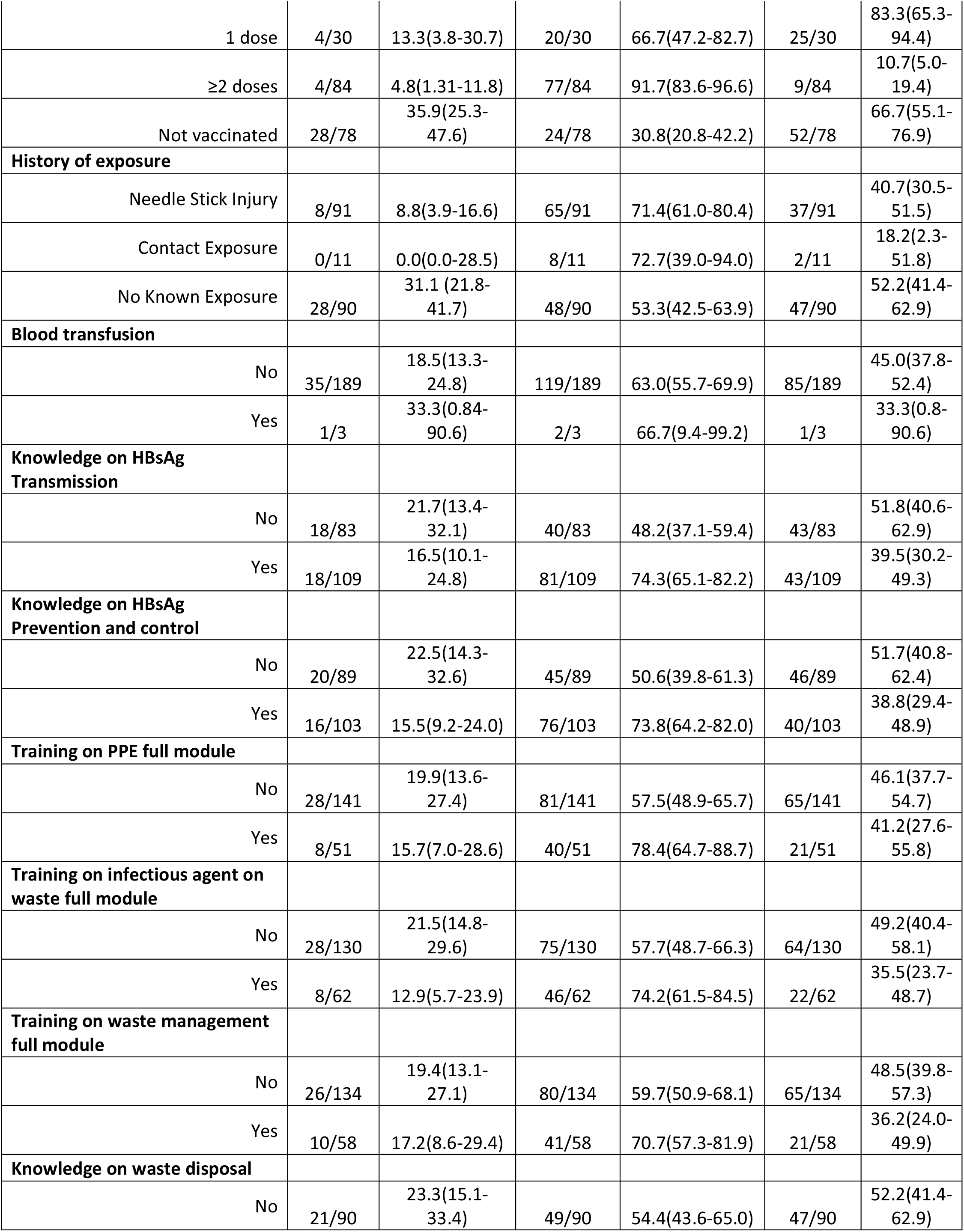
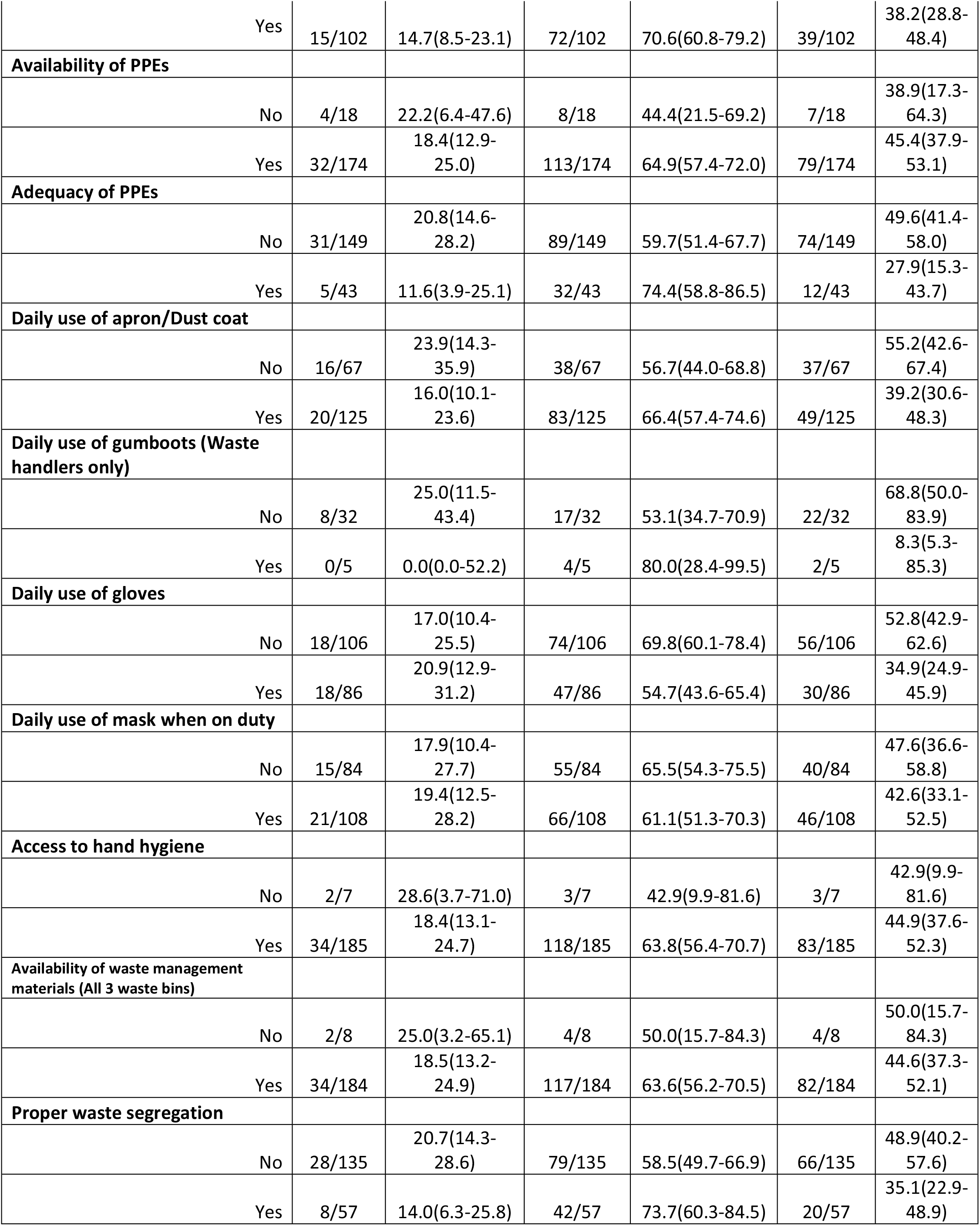
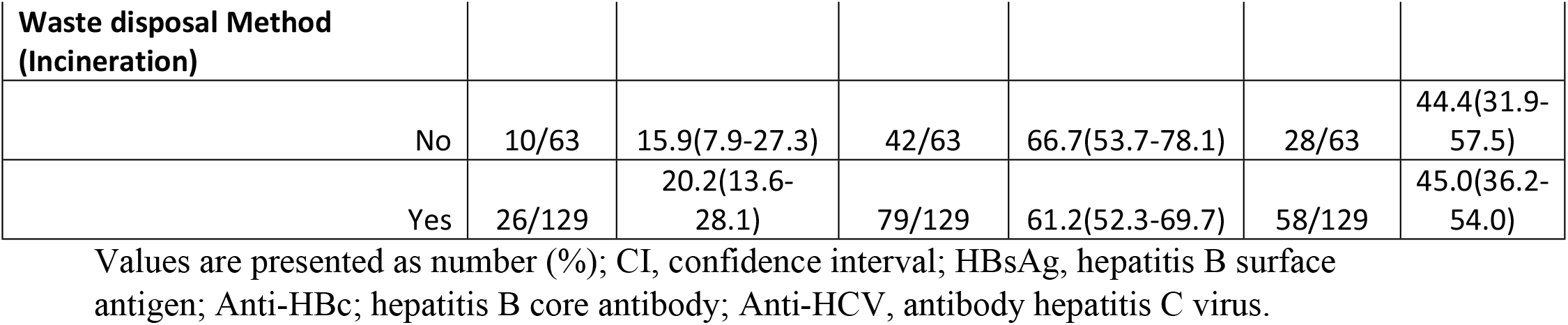
Prevalence of hepatitis B virus markers among HCW in Kisumu County, 2020 (N= 192)

| Characteristic | Prevalence of HBsAg Biomarker | Prevalence of Anti-HBs Biomarker | Prevalence of Anti-HBc Biomarker |
| --- | --- | --- | --- |

|  | N(%) | % (95%CI) | N(%) | % (95%CI) | N(%) | % (95%CI) |
| --- | --- | --- | --- | --- | --- | --- |
| Overall | 36/192 | 18.8(13.5-25.0) | 121/192 | 63.0(55.8-69.9) | 86/192 | 44.8(37.6-52.1) |
| **Carder** |  |  |  |  |  |  |
| Doctors | 2/8 | 25.0(3.19-65.1) | 5/8 | 62.5(24.5-91.5) | 3/8 | 37.5(8.5-75.5) |
| Clinical Officer | 6/29 | 20.7(8-39.7) | 19/29 | 65.5(45.7-82.1) | 14/29 | 48.3(29.5-67.5) |
| Nursing officer | 8/39 | 20.5(9.3-36.5) | 25/39 | 64.1(47.2-78.8) | 15/39 | 38.5(23.4-55.4) |
| Laboratory technologist | 7/55 | 12.7(5.3-24.5) | 41/55 | 74.6(61.0-85.3) | 18/55 | 32.7(20.7-46.7) |
| Mortuary attendance | 0/7 | 0.0(0-41.0) | 4/7 | 57.1(18.4-90.1) | 4/7 | 57.1(18.4-90.1) |
| HTS counsellor | 5/17 | 29.4(10.3-56) | 6/17 | 35.3(14.2-61.7) | 8/17 | 47.1(23.0-72.2) |
| Waste handlers | 8/37 | 21.6(9.8-38.2) | 21/37 | 56.8(39.5-72.9) | 24/37 | 64.9(47.5-79.8) |
| **Sex** |  |  |  |  |  |  |
| Male | 12/100 | 12(6.4-20.0) | 69/100 | 69.0(59.0-77.9) | 46/100 | 46.0(36.0-56.3) |
| Female | 24/92 | 26.1(17.5-36.3) | 52/92 | 56.5(45.8-66.8) | 40/92 | 43.5(33.2-54.2) |
| **Age(Years)** |  |  |  |  |  |  |
| 20-29 | 10/65 | 15.4(7.6-26.5) | 41/65 | 63.1(50.2-74.7) | 25/65 | 38.5(26.7-51.4) |
| 30-39 | 15/83 | 18.1(10.5-28.1) | 56/83 | 67.5(56.3-77.4) | 35/83 | 42.2(31.4-53.5) |
| 40-49 | 6/23 | 26.1(10.2-48.4) | 11/23 | 47.8(26.8-69.4) | 14/23 | 60.9(38.5-80.3) |
| ≥50 | 5/21 | 23.8(8.2-47.2) | 13/21 | 61.9(38.4-85.1) | 12/21 | 57.1(34.0-78.2) |
| **Marital status** |  |  |  |  |  |  |
| Single | 7/37 | 18.9(8.0-35.2) | 20/37 | 54.1(36.9-70.5) | 15/37 | 40.5(24.8-57.9) |
| Married | 28/151 | 18.5(12.7-25.7) | 98/151 | 64.9(56.7-72.5) | 69/151 | 45.7(37.6-54.0) |
| Widowed/Separated/Divorced | 1/4 | 25(0.6-80.6) | 3/4 | 75.0(19.4-99.4) | 2/4 | 50.0(6.8-93.2) |
| **Years of service(Years)** |  |  |  |  |  |  |
| <1 | 3/8 | 37.5(8.5-75.5) | 4/8 | 50.0(15.7-84.3) | 6/8 | 75.0(34.9-96.8) |
| 1-5 | 13/85 | 15.3(8.4-24.7) | 51/85 | 60.0(48.8-70.5) | 31/85 | 36.5(26.3-47.6) |
| 6-10 | 10/51 | 19.6(9.8-33.1) | 38/51 | 74.5(60.4-85.7) | 26/51 | 51.0(36.6-65.3) |
| >10 | 10/48 | 20.8(10.5-35.0) | 28/48 | 58.3(43.2-72.4) | 23/48 | 47.9(33.3-62.8) |
| **HBV Vaccine uptake** |  |  |  |  |  |  |
| 1 dose | 4/30 | 13.3(3.8-30.7) | 20/30 | 66.7(47.2-82.7) | 25/30 | 83.3(65.3-94.4) |
| ≥2 doses | 4/84 | 4.8(1.31-11.8) | 77/84 | 91.7(83.6-96.6) | 9/84 | 10.7(5.0-19.4) |
| Not vaccinated | 28/78 | 35.9(25.3-47.6) | 24/78 | 30.8(20.8-42.2) | 52/78 | 66.7(55.1-76.9) |
| **History of exposure** |  |  |  |  |  |  |
| Needle Stick Injury | 8/91 | 8.8(3.9-16.6) | 65/91 | 71.4(61.0-80.4) | 37/91 | 40.7(30.5-51.5) |
| Contact Exposure | 0/11 | 0.0(0.0-28.5) | 8/11 | 72.7(39.0-94.0) | 2/11 | 18.2(2.3-51.8) |
| No Known Exposure | 28/90 | 31.1 (21.8-41.7) | 48/90 | 53.3(42.5-63.9) | 47/90 | 52.2(41.4-62.9) |
| **Blood transfusion** |  |  |  |  |  |  |
| No | 35/189 | 18.5(13.3-24.8) | 119/189 | 63.0(55.7-69.9) | 85/189 | 45.0(37.8-52.4) |
| Yes | 1/3 | 33.3(0.84-90.6) | 2/3 | 66.7(9.4-99.2) | 1/3 | 33.3(0.8-90.6) |
| **Knowledge on HBsAg Transmission** |  |  |  |  |  |  |
| No | 18/83 | 21.7(13.4-32.1) | 40/83 | 48.2(37.1-59.4) | 43/83 | 51.8(40.6-62.9) |
| Yes | 18/109 | 16.5(10.1-24.8) | 81/109 | 74.3(65.1-82.2) | 43/109 | 39.5(30.2-49.3) |
| **Knowledge on HBsAg Prevention and control** |  |  |  |  |  |  |
| No | 20/89 | 22.5(14.3-32.6) | 45/89 | 50.6(39.8-61.3) | 46/89 | 51.7(40.8-62.4) |
| Yes | 16/103 | 15.5(9.2-24.0) | 76/103 | 73.8(64.2-82.0) | 40/103 | 38.8(29.4-48.9) |
| **Training on PPE full module** |  |  |  |  |  |  |
| No | 28/141 | 19.9(13.6-27.4) | 81/141 | 57.5(48.9-65.7) | 65/141 | 46.1(37.7-54.7) |
| Yes | 8/51 | 15.7(7.0-28.6) | 40/51 | 78.4(64.7-88.7) | 21/51 | 41.2(27.6-55.8) |
| **Training on infectious agent on waste full module** |  |  |  |  |  |  |
| No | 28/130 | 21.5(14.8-29.6) | 75/130 | 57.7(48.7-66.3) | 64/130 | 49.2(40.4-58.1) |
| Yes | 8/62 | 12.9(5.7-23.9) | 46/62 | 74.2(61.5-84.5) | 22/62 | 35.5(23.7-48.7) |
| **Training on waste management full module** |  |  |  |  |  |  |
| No | 26/134 | 19.4(13.1-27.1) | 80/134 | 59.7(50.9-68.1) | 65/134 | 48.5(39.8-57.3) |
| Yes | 10/58 | 17.2(8.6-29.4) | 41/58 | 70.7(57.3-81.9) | 21/58 | 36.2(24.0-49.9) |
| **Knowledge on waste disposal** |  |  |  |  |  |  |
| No | 21/90 | 23.3(15.1-33.4) | 49/90 | 54.4(43.6-65.0) | 47/90 | 52.2(41.4-62.9) |
| Yes | 15/102 | 14.7(8.5-23.1) | 72/102 | 70.6(60.8-79.2) | 39/102 | 38.2(28.8-48.4) |
| **Availability of PPEs** |  |  |  |  |  |  |
| No | 4/18 | 22.2(6.4-47.6) | 8/18 | 44.4(21.5-69.2) | 7/18 | 38.9(17.3-64.3) |
| Yes | 32/174 | 18.4(12.9-25.0) | 113/174 | 64.9(57.4-72.0) | 79/174 | 45.4(37.9-53.1) |
| **Adequacy of PPEs** |  |  |  |  |  |  |
| No | 31/149 | 20.8(14.6-28.2) | 89/149 | 59.7(51.4-67.7) | 74/149 | 49.6(41.4-58.0) |
| Yes | 5/43 | 11.6(3.9-25.1) | 32/43 | 74.4(58.8-86.5) | 12/43 | 27.9(15.3-43.7) |
| **Daily use of apron/Dust coat** |  |  |  |  |  |  |
| No | 16/67 | 23.9(14.3-35.9) | 38/67 | 56.7(44.0-68.8) | 37/67 | 55.2(42.6-67.4) |
| Yes | 20/125 | 16.0(10.1-23.6) | 83/125 | 66.4(57.4-74.6) | 49/125 | 39.2(30.6-48.3) |
| **Daily use of gumboots (Waste handlers only)** |  |  |  |  |  |  |
| No | 8/32 | 25.0(11.5-43.4) | 17/32 | 53.1(34.7-70.9) | 22/32 | 68.8(50.0-83.9) |
| Yes | 0/5 | 0.0(0.0-52.2) | 4/5 | 80.0(28.4-99.5) | 2/5 | 8.3(5.3-85.3) |
| **Daily use of gloves** |  |  |  |  |  |  |
| No | 18/106 | 17.0(10.4-25.5) | 74/106 | 69.8(60.1-78.4) | 56/106 | 52.8(42.9-62.6) |
| Yes | 18/86 | 20.9(12.9-31.2) | 47/86 | 54.7(43.6-65.4) | 30/86 | 34.9(24.9-45.9) |
| **Daily use of mask when on duty** |  |  |  |  |  |  |
| No | 15/84 | 17.9(10.4-27.7) | 55/84 | 65.5(54.3-75.5) | 40/84 | 47.6(36.6-58.8) |
| Yes | 21/108 | 19.4(12.5-28.2) | 66/108 | 61.1(51.3-70.3) | 46/108 | 42.6(33.1-52.5) |
| **Access to hand hygiene** |  |  |  |  |  |  |
| No | 2/7 | 28.6(3.7-71.0) | 3/7 | 42.9(9.9-81.6) | 3/7 | 42.9(9.9-81.6) |
| Yes | 34/185 | 18.4(13.1-24.7) | 118/185 | 63.8(56.4-70.7) | 83/185 | 44.9(37.6-52.3) |
| **Availability of waste management materials (All 3 waste bins)** |  |  |  |  |  |  |
| No | 2/8 | 25.0(3.2-65.1) | 4/8 | 50.0(15.7-84.3) | 4/8 | 50.0(15.7-84.3) |
| Yes | 34/184 | 18.5(13.2-24.9) | 117/184 | 63.6(56.2-70.5) | 82/184 | 44.6(37.3-52.1) |
| **Proper waste segregation** |  |  |  |  |  |  |
| No | 28/135 | 20.7(14.3-28.6) | 79/135 | 58.5(49.7-66.9) | 66/135 | 48.9(40.2-57.6) |
| Yes | 8/57 | 14.0(6.3-25.8) | 42/57 | 73.7(60.3-84.5) | 20/57 | 35.1(22.9-48.9) |

| Waste disposal Method<br>(Incineration) |  |  |  |  |  |  |
| --- | --- | --- | --- | --- | --- | --- |
| No | 10/63 | 15.9(7.9-27.3) | 42/63 | 66.7(53.7-78.1) | 28/63 | 44.4(31.9-57.5) |
| Yes | 26/129 | 20.2(13.6-28.1) | 79/129 | 61.2(52.3-69.7) | 58/129 | 45.0(36.2-54.0) |

Values are presented as number (%); CI, confidence interval; HBsAg, hepatitis B surface antigen; Anti-HBc, hepatitis B core antibody; Anti-HCV, antibody hepatitis C virus.

**Table 3:** Factors associated with current HBV infection among HCW in Kisumu County.

|  |  |  |  |  |  |  |
| --- | --- | --- | --- | --- | --- | --- |
| Characteristic | HWC (%) | N(%) | OR (95% CI) | p-value | aOR(95% CI) | p-value |
| Overall | N(%) | 36 (18.8) |  |  |  |  |
| **Cadre** |  |  |  |  |  |  |
| Doctors | 8(4.2) | 2(25.0) | 1 |  |  |  |
| Clinical Officer | 29(15.1) | 6(20.7) | 0.78(0.12-4.90) | 0.793 |  |  |
| Nursing officer | 39(20.3) | 8(20.5) | 0.77(0.13-4.59) | 0.778 |  |  |
| Laboratory technologist | 55(28.7) | 7(12.7) | 0.44(0.07-2.61) | 0.364 |  |  |
| Mortuary attendance | 7(3.7) | 0(0.0) | - |  |  |  |
| HTS counsellor | 17(9.9) | 5(29.4) | 1.25(0.19-8.44) | 0.819 |  |  |
| Waste handlers | 37(19.3) | 8(21.6) | 0.83(0.14-4.91) | 0.835 |  |  |
| **Sex** |  |  |  |  |  |  |
| Male | 92(47.9) | 12(12.0) | 1 |  | 1 |  |
| Female | 100(52.1) | 24(26.1) | 2.59(1.21-5.54) | 0.014 | 3.22(1.06-9.75) | 0.039 |
| **Age(years)** |  |  |  |  |  |  |
| 20-29 | 65(33.9) | 10(15.4) | 1 |  |  |  |
| 30-39 | 83(43.2) | 15(18.1) | 1.21(0.51-2.91) | 0.665 |  |  |
| 40-49 | 23(12.0) | 6(26.1) | 1.94(0.62-6.12) | 0.258 |  |  |
| ≥50 | 21(10.9) | 5(23.8) | 1.72(0.51<br>-5.76) | 0.38 |  |  |
| **Marital status** |  |  |  |  |  |  |
| Single | 37(19.3) | 7(18.9) | 1 |  |  |  |
| Married | 151(78.7<br>) | 28(18.5) | 0.98(0.39<br>-2.45) | 0.958 |  |  |
| Widowed/Separated/Divorced | 4(2.1) | 1(25.0) | 1.42(0.13<br>-15.87) | 0.772 |  |  |
| **Years of service(years)** |  |  |  |  |  |  |
| <1 | 8(4.2) | 3(37.5) | 1 |  | 1 |  |
| 1-5 | 85(44.3) | 13(15.3) | 0.30(0.06<br>-1.42) | 0.128 | 0.56(0.08-3.98) | 0.56 |
| 6-10 | 51(26.6) | 10(19.6) | 0.41(0.08<br>-1.99) | 0.267 | 1.89(0.17-21.55) | 0.607 |
| >10 | 48(25.0) | 10(20.8) | 0.44(0.09<br>-2.15) | 0.31 | 1.61(0.1-25.91) | 0.738 |
| **HBV Vaccine uptake** |  |  |  |  |  |  |
| Not vaccinated | 78(40.6) | 28(35.9) | 1 |  | 1 |  |
| 1 dose | 30(15.6) | 4(13.3) | 0.27 | 0.028 | 0.31(0.06-1.54) | 0.153 |
| ≥2 doses | 84(43.8) | 4(4.8) | 0.09 | <0.000<br>1 | 0.05(0.01-0.20) | <0.000<br>1 |
| **History of exposure** |  |  |  |  |  |  |
| Needle Stick Injury | 91(47.4) | 8(8.8) | 1 |  | 1 |  |
| Contact Exposure | 11(5.7) | 0(0.0) | 1(-) |  | 1(-) |  |
| No Known Exposure | 90(46.9) | 28(31.1) | 4.69(2.00<br>-10.98) | <0.001 | 5.37(1.81-15.92) | <0.00<br>1 |
| **Blood transfusion** |  |  |  |  |  |  |
| No | 189(98.4<br>) | 35(18.5) | 1 |  |  |  |
| Yes | 3(1.6) | 1(33.3) | 2.20(0.19<br>-24.95) | 0.525 |  |  |
| **Knowledge on HBsAg Transmission** |  |  |  |  |  |  |
| No | 83(43.2) | 18(21.7) | 1 |  |  |  |
| Yes | 109(56.8<br>) | 18(16.5) | 0.71(0.35<br>-1.48) | 0.364 |  |  |
| **Knowledge on HBsAg Prevention and control** |  |  |  |  |  |  |
| No | 89(46.4) | 20(22.5) | 1 |  |  |  |
| Yes | 103(53.7<br>) | 16(15.5) | 0.63(0.31<br>-1.32) | 0.222 |  |  |
| **Training on PPE full module** |  |  |  |  |  |  |
| No | 141(26.6<br>) | 28(19.9) | 1 |  |  |  |
| Yes | 51(73.4) | 8(15.7) | 0.75(0.32<br>-1.78) | 0.514 |  |  |
| **Training on infectious agent on waste full module** |  |  |  |  |  |  |
| No | 130(67.7) | 28(21.5) | 1 |  | 1 |  |
| Yes | 62(32.3) | 8(12.9) | 0.54(0.23<br>-1.27) | 0.156 | 1.53(0.31-7.57) | 0.604 |
| **Training on waste management full module** |  |  |  |  |  |  |
| No | 134(69.8) | 26(19.4) | 1 |  |  |  |
| Yes | 58(30.2) | 10(17.2) | 0.87(0.39<br>-1.93) | 0.725 |  |  |
| **Knowledge on waste disposal** |  |  |  |  |  |  |
| No | 90(46.9) | 21(23.3) | 1 |  | 1 |  |
| Yes | 102(53.1) | 15(14.7) | 0.57(0.27<br>-1.18) | 0.129 | 0.49(0.13-1.86) | 0.297 |
| **Availability of PPEs** |  |  |  |  |  |  |
| No | 18(9.4) | 4(22.2) | 1 |  |  |  |
| Yes | 174(90.6) | 32(18.4) | 0.79(0.24<br>-2.56) | 0.692 |  |  |
| **Adequacy of PPEs** |  |  |  |  |  |  |
| No | 149(77.6) | 27(19.6) | 1 |  | 1 |  |
| Yes | 43(22.4) | 5(11.6) | 0.50(0.18<br>-1.38) | 0.181 | 0.47(0.13-1.72) | 0.256 |
| **Daily use of apron/Dust coat** |  |  |  |  |  |  |
| No | 67(34.9) | 16(23.9) | 1 |  | 1 |  |
| Yes | 125(65.1) | 20(16.0) | 0.61(0.29<br>-1.27) | 0.185 | 0.57(0.20-1.64) | 0.297 |
| **Daily use of gumboots (Waste handlers only)** |  |  |  |  |  |  |
| No | 32(86.5) | 5(13.5) | 1 |  |  |  |
| Yes | 5(13.5) | 32(86.5) | 1(-) | - |  |  |
| **Daily use of gloves** |  |  |  |  |  |  |
| No | 106(55.2) | 18(17.0) | 1 |  |  |  |
| Yes | 86(44.8) | 18(20.9) | 1.29(0.63<br>-2.67) | 0.486 |  |  |
| **Daily use of mask when on duty** |  |  |  |  |  |  |
| No | 84(43.8) | 15(17.9) | 1 |  |  |  |
| Yes | 108(56.3<br>) | 21(19.4) | 1.11(0.53<br>-2.31) | 0.78 |  |  |
| **Access to hand hygiene** |  |  |  |  |  |  |
| No | 7(3.7) | 2(28.6) | 1 |  |  |  |
| Yes | 185(96.4<br>) | 34(18.4) | 0.56(0.10<br>-3.02) | 0.503 |  |  |
| **Availability of waste management materials (All 3 waste bins)** |  |  |  |  |  |  |
| No | 8(4.2) | 2(25.0) | 1 |  |  |  |
| Yes | 184(95.8<br>) | 34(18.5) | 0.68(0.13<br>-3.52) | 0.645 |  |  |
| **Proper waste segregation** |  |  |  |  |  |  |
| No | 135(70.3<br>) | 28(20.7) | 1 |  |  |  |
| Yes | 57(29.7) | 8(14.0) | 0.62(0.27<br>-1.47) | 0.28 |  |  |
| **Waste disposal Method (Incineration)** |  |  |  |  |  |  |
| No | 63(32.8) | 10(15.9) | 1 |  |  |  |
| Yes | 129(67.2<br>) | 26(20.2) | 1.34(0.60<br>-2.98) | 0.476 |  |  |

Values are presented as number (%); OR, odds ratio; aOR, adjusted odds ratio; 95% CI, confidence interval.

Anti HBc prevalence was highest among HCW with one dose of HBV vaccine 83.3% (95% CI: 65.3-94.4), those with less than one year in service 75.0% (95% CI: 34.9-96.8), waste handlers not using gumboots 68.8% (95% CI: 50.0-83.9) and HBV unvaccinated HCW 66.7% (95% CI: 55.1-76.9).

There is moderate HBV immunity or recovery level among HCW, the carders with highest anti HBs positivity were laboratory scientist 74.6% (95% CI: 61.0-85.3), clinical officers 65.5% (95% CI:45.7-82.1) and Nursing officers 64.1% (95% CI: 47.2-78.8). HTS counselors had the lowest immunity or recovery level at 35.3% (95% CI: 14.2-61.7).

### HBV infection by Vaccine uptake

The prevalence of HBV was highest among healthcare workers who had not received any dose of HBV vaccine (at 35.9%), those who received one dose of HBV vaccination had a prevalence of 13.3%, while those who received two or more doses of HBV vaccination had a prevalence of 4.8% (Figure 1). Notably, none of the 69 HCWs who reported receiving all the three required doses of HBV vaccination were detected with HBV infection.

**Figure 1:**
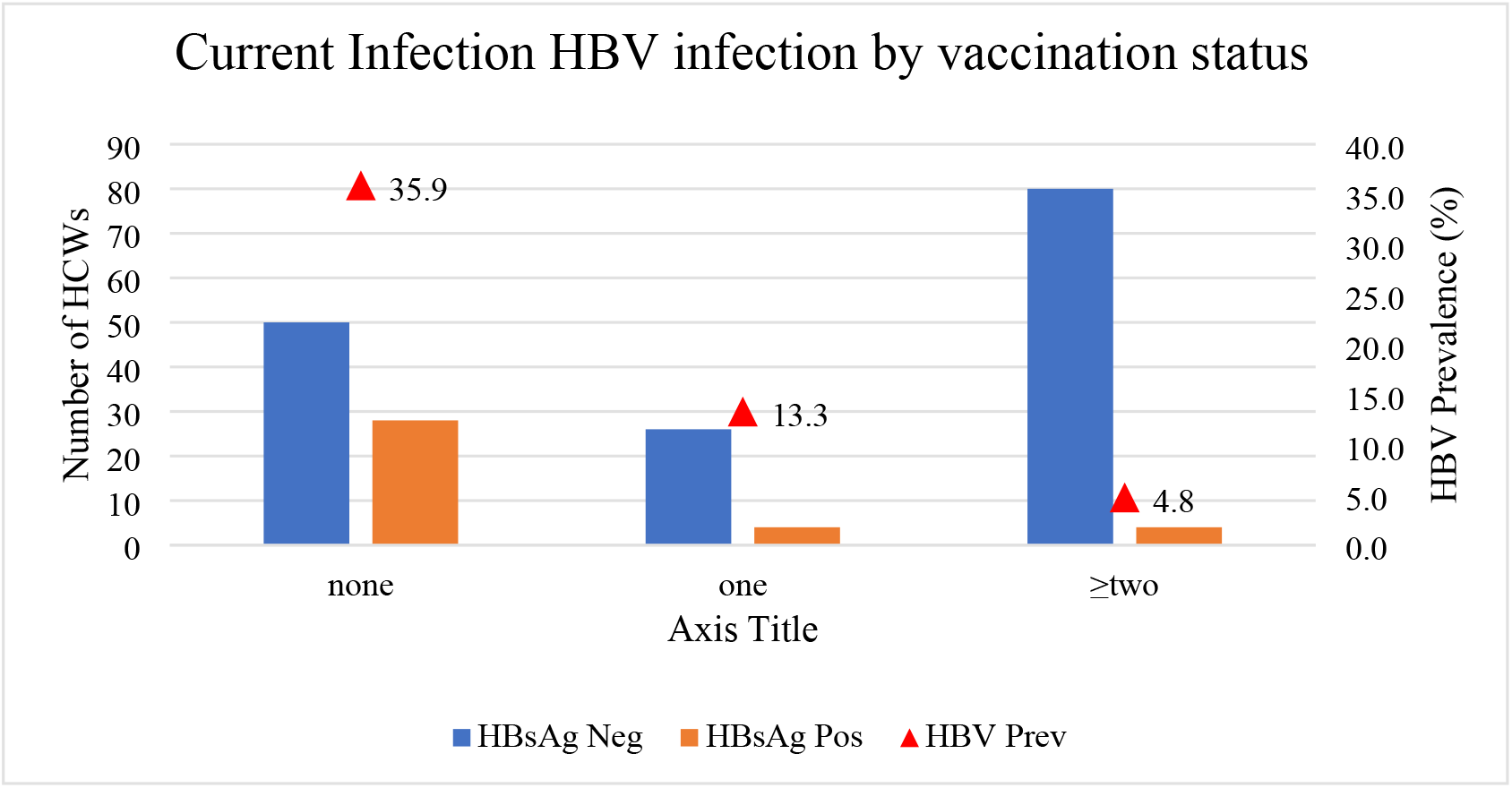
HBV infection by Vaccination status.

### Factors associated with Current HBV infection

Table3 shows that female HCW were more likely to have current HBV infection compared to their male counterparts (aOR, 3.22; 95% CI, 1.06-9.75, p-value< 0.05). Additionally, HCW without a history of known exposure had increased odds of current HBV infection compared to those with a previous needle stick injury (aOR, 5.37; 95% CI, 1.81-15.92, p-value< 0.001). However, HCW who reported receiving ≥2 doses of HBV vaccination had reduced likelihood of current HBV infection. (aOR, 0.05; 95% CI, 0.01-0.20, p-value <0.001) respectively. None of the other sociodemographic characteristics were associated with current infection of HBV among HCW.

### Factors associated with lifetime exposure to HBV infection among HCW in Kisumu County, 2020

In logistic regression analysis Table 4, HCW who received a single dose of HBV vaccination had increased likelihood of lifetime exposure to HBV infection compared to HCW without history of vaccination (aOR, 6.25; 95% CI, 1.29-30.30, p-value<0.05). Conversely, HCW who reported receiving ≥2 doses of HBV vaccination had reduced likelihood of lifetime exposure to HBV infection compared to those without HBV vaccination (aOR, 0.03; 95% CI, 0.01-0.10, p-value= <0.0001) HCW who reported having knowledge on HBsAg transmission had higher odds of lifetime exposure to HBV infection compared to their counterparts without knowledge on HBV transmission (aOR, 7.97; 95% CI, 2.10-153.39, p-value<0.01). None of the other sociodemographic characteristics were significantly associated with current infection of HBV in HCW (Table 4).

**Table 4:** Factors associated with lifetime exposure to HBV infection among Health care workers in Kisumu County, 2020.

| Characteristics | HCW (%) | N(%) | OR (95%CI) | p-value | aOR (95% CI) | p-value |
| --- | --- | --- | --- | --- | --- | --- |
| **Sex** |  |  |  |  |  |  |
| Female | 92(47.9) | 40(43.5) | 1 |  |  |  |
| Male | 100(52.1) | 46(46.0) | 1.11(0.63-1.96) | 0.726 |  |  |
| **Age** |  |  |  |  |  |  |
| 20-29 | 65(33.9) | 25(38.5) | 1 |  | 1 |  |
| 30-39 | 83(43.2) | 35(42.2) | 1.17(0.60-2.26) | 0.649 | 1.18(0.37-3.77) | 0.784 |
| 40-49 | 23(12.0) | 14(60.9) | 2.49(0.94-6.60) | 0.067 | 1.77(0.28-11.37) | 0.547 |
| 50-59 | 18(9.4) | 10(55.6) | 2.00(0.70-5.75) | 0.198 | 3.41(0.33-34.81) | 0.301 |
| $\geq 60$ | 3(1.6) | 2(66.7) | 3.2(0.28-37.15) | 0.352 | 0.27(0.01-7.98) | 0.452 |
| **Marital status** |  |  |  |  |  |  |
| Single | 37(19.3) | 15(40.5) | 1 |  |  |  |
| Married | 151(78.7) | 69(45.7) | 1.23(0.59-2.56) | 0.572 |  |  |
| Widowed/Separated/Divorced | 4(2.1) | 2(50.0) | 1.47(0.19-11.59) | 0.716 |  |  |
| **Year of service** |  |  |  |  |  |  |
| Less than 1 year | 8(4.2) | 6(75.0) | 1 |  | 1 |  |
| 1-5 years | 85(44.3) | 31(36.5) | 0.19(0.04-1.01) | **0.051** | 0.13(0.01-2.79) | 0.193 |
| 6-10 years | 51(26.6) | 26(51.0) | 0.35(0.06-1.88) | 0.22 | 0.57(0.02-15.34) | 0.741 |
| More than 10 years | 48(25.0) | 23(47.9) | 0.31(0.06-1.67) | 0.172 | 0.31(0.01-9.82) | 0.509 |
| **Carder** |  |  |  |  |  |  |
| Doctors | 8(4.2) | 3(37.5) | 1 |  | 1 |  |
| Clinical Officer | 29(15.1) | 14(48.3) | 1.56(0.31-7.75) | 0.59 | 0.72(0.05-10.26) | 0.811 |
| Nursing officer | 39(20.3) | 15(38.5) | 1.04(0.22-5.01) | 0.959 | 0.49(0.04-6.59) | 0.589 |
| Laboratory technologist | 55(28.7) | 18(32.7) | 0.81(0.17-3.78) | 0.789 | 0.47(0.05-4.87) | 0.527 |
| Mortuary attendance | 7(3.7) | 4(57.1) | 2.22(0.28-17.63) | 0.45 | 0.39(0.02-8.56) | 0.578 |
| HTS counsellor | 17(9.9) | 8(47.1) | 1.48(0.27-8.27) | 0.654 | 0.73(0.05-9.71) | 0.811 |
| Waste handlers | 37(19.3) | 24(64.9) | 3.08(0.63-14.98) | 0.164 | 2.00(0.17-22.84) | 0.578 |
| **HBV Vaccine uptake** |  |  |  |  |  |  |
| Not vaccinated | 78(40.6) | 52(66.7) | 1 |  | 1 |  |
| 1 dose | 30(15.6) | 25(83.3) | 2.5(0.86-7.28) | 0.093 | 6.25(1.29-30.30) | 0.023 |
| ≥2 doses | 84(43.8) | 9(10.7) | 0.06(0.03-0.14) | <0.0001 | 0.03(0.01-0.10) | <0.0001 |
| **History of exposure** |  |  |  |  |  |  |
| Needle Stick Injury | 91(47.4) | 37(40.7) | 1 |  | 1 |  |
| Contact Exposure | 11(5.7) | 2(18.2) | 0.32(0.07-1.59) | 0.165 | 0.10(0.01-1.40) | 0.087 |
| No Known Exposure | 90(46.9) | 47(52.2) | 1.59(0.89-2.87) | 0.12 | 3.55(1.34-9.43) | 0.011 |
| **Blood transfusion** |  |  |  |  |  |  |
| No | 189(98.4) | 85(45.0) | 1 |  |  |  |
| Yes | 3(1.6) | 1(33.3) | 0.61(0.05-6.86) | 0.69 |  |  |
| **Knowledge on HBsAg Transmission** |  |  |  |  |  |  |
| No | 83(43.2) | 43(51.8) | 1 |  | 1 |  |
| Yes | 109(56.8) | 43(39.5) | 0.61(0.34-1.08) | 0.089 | 17.97(2.10-153.39) | 0.008 |
| **Knowledge on HBsAg Prevention and control** |  |  |  |  |  |  |
| No | 89(46.4) | 46(51.7) | 1 |  | 1 |  |
| Yes | 103(53.7) | 40(38.8) | 0.59(0.33-1.05) | 0.075 | 0.34(0.06-2.12) | 0.25 |
| **Training on PPE full module** |  |  |  |  |  |  |
| No | 141(26.6) | 65(46.1) | 1 |  |  |  |
| Yes | 51(73.4) | 21(41.2) | 0.82(0.43-1.57) | 0.545 |  |  |
| **Training on infectious agent on waste full module** |  |  |  |  |  |  |
| No | 130(67.7) | 64(49.2) | 1 |  | 1 |  |
| Yes | 62(32.3) | 22(35.5) | 0.57(0.30-1.06) | 0.075 | 0.65(0.10-4.21) | 0.652 |
| **Training on waste management full module** |  |  |  |  |  |  |
| No | 134(69.8) | 65(48.5) | 1 |  | 1 |  |
| Yes | 58(30.2) | 21(36.2) | 0.60(0.32-1.14) | 0.117 | 2.37(0.32-17.28) | 0.395 |
| **Knowledge on waste disposal** |  |  |  |  |  |  |
| No | 90(46.9) | 47(52.2) | 1 |  | 1 |  |
| Yes | 102(53.1) | 39(38.2) | 0.57(0.32-1.01) | 0.053 | 0.41(0.11-1.47) | 0.17 |
| **Availability of PPEs** |  |  |  |  |  |  |
| No | 18(9.4) | 7(38.9) | 1 |  |  |  |
| Yes | 174(90.6) | 79(45.4) | 1.31(0.48-3.53) | 0.598 |  |  |
| **Adequacy of PPEs** |  |  |  |  |  |  |
| No | 149(77.6) | 74(49.7) | 1 |  | 1 |  |
| Yes | 43(22.4) | 12(27.9) | 0.39(0.19-0.82) | 0.013 | 0.49(0.16-1.56) | 0.228 |
| **Daily use of apron/Dust coat** |  |  |  |  |  |  |
| No | 67(34.9) | 37(55.2) | 1 |  | 1 |  |
| Yes | 125(65.1) | 49(39.2) | 0.52(0.29-0.95) | 0.034 | 0.73(0.26-2.03) | 0.549 |
| **Daily use of gumboots (Waste handlers only)** |  |  |  |  |  |  |
| No | 32(86.5) | 22(68.8) | 1 |  |  |  |
| Yes | 5(13.5) | 2(8.3) | 0.82(0.13-5.01) | 0.827 |  |  |
| **Daily use of gloves** |  |  |  |  |  |  |
| No | 106(55.2) | 56(52.8) | 1 |  | 1 |  |
| Yes | 86(44.8) | 30(34.9) | 0.48(0.27-0.86) | 0.013 | 0.40(0.16-1.01) | 0.052 |
| **Daily use of mask when on duty** |  |  |  |  |  |  |
| No | 84(43.8) | 40(47.6) | 1 |  |  |  |
| Yes | 108(56.3) | 46(42.6) | 0.82(0.56-1.45) | 0.487 |  |  |
| **Access to hand hygiene** |  |  |  |  |  |  |
| No | 7(3.7) | 3(42.9) | 1 |  |  |  |
| Yes | 185(96.4) | 83(44.9) | 1.08(0.24-4.98) | 0.917 |  |  |
| **Availability of waste management materials (All 3 waste bins)** |  |  |  |  |  |  |
| No | 8(4.2) | 4(50.0) | 1 |  |  |  |
| Yes | 184(95.8) | 82(44.6) | 0.80(0.20-3.31) | 0.763 |  |  |
| **Proper waste segregation** |  |  |  |  |  |  |
| No | 135(70.3) | 66(48.9) | 1 |  | 1 |  |
| Yes | 57(29.7) | 20(35.1) | 0.57(0.30-1.07) | 0.081 | 0.94(0.28-3.19) | 0.991 |
| **Waste disposal Method (Incineration)** |  |  |  |  |  |  |
| No | 63(32.8) | 28(44.4) | 1 |  |  |  |
| Yes | 129(67.2) | 58(45.0) | 1.02(0.56-1.87) | 0.946 |  |  |

Values are presented as number (%); OR odds ratio; aOR, adjusted odds ratio; CI, confidence interval.

## Discussion

Indeed the burden of Hospital acquired infection like Hepatitis B infection is high in developing sub-Saharan African countries like Kenya (13). Despite the availability of guidelines and treatment options, occupational risks related to hepatitis virus exposure is still a major concern for those who handle hospital waste like health care workers (14). This study documented high prevalence and lifetime exposure to HBV infection in Kisumu county, Kenya at 18.8% and 44.8% respectively, the subpopulations with the highest HBV infection are: HCW who had worked for less than 1 year at 37.5%,HBV unvaccinated HCW at 35.9%, HCW with previous history of blood transfusion at 33.3% and HIV testing counselor at 29.4%. The HBV prevalence among HCW in this study is higher compared to 2.7% prevalence in general population(15). This prevalence is higher than what was found in other studies both in Kenya and Africa : pooled prevalence in Africa 6.8%, Kenya 4.0 %, southern Ethiopia 1.3%, north west Ethiopia 6.0 %, Tripoli Libya 2.3% and 6.3% in Addis Ababa, (2–5, 14, 16). A study in Kisumu, Siaya and Homabay county found that the prevalence of HBV among adolescent was 3.4% while dual infection of HBV and HIV in Kisumu county hospital, one of our study site was 47% among patient presenting with jaundice in the clinic (17, 18). The high prevalence in this study could be attributed to increased HBV infection risk of exposure on HCW as highlighted in this study where 53.7% of HCW has had either needle stick or contact exposure, inadequate trainings, observed knowledge gaps, poor infection prevention infrastructure, low Hepatitis B virus vaccine coverage and low-adherence universal infection prevention and control measures (consistent use of PPE, proper waste segregation and disposal) as shown in table 1. Low training coverage on waste management, infectious agent on waste and inadequacy of PPE at 30.2%,32.2%, 22.4% respectively could be the cause to high needle stick injury, contact exposure and poor adherence to standard precautions. The findings on inadequate HCW capacity building, poor adherence on IPC standard and additional precautions are in concurrence with other studies which pointed to poor waste management and lack of training and capacity building to staff (16, 19, 20). Health care system administrators should ensure adequate PPEs are available, accessible and properly used by HCW, IPC trainings should be done to all health care workers with provision of annual refresher.

Hepatitis B surface antibody (anti-HBs) prevalence in this study was 63.0%, these were HCW who have recovered from infection or immunised HBV. Anti HBS positivity was highest in laboratory technologists 74.6%, clinical officers 65.5%, and Nursing officers 64.1%. Laboratory technologists were the highest immunized carder, this may be attributed to the implementation of laboratory quality management system and international organization for standardization ISO 15189 which required that all laboratory personnel must be vaccinated against blood borne pathogens (21). The low HBV immunity rate and non-completion of vaccine doses is comparable to prior studies done in the region showing low HBV vaccine uptake and non-completion of vaccine dosses(2, 16). The benefit vaccination as HBV infection prevention measure has been documented, this study also found HCW who are fully vaccinated had no lifetime exposure while unvaccinated HCW had highest 61% lifeline exposure to HBV infection (22).Findings in this study highlights the need to capacity build HCW on the benefits of getting full vaccine dose and need to avail the vaccine for all health care workforce.. significant risk of lifetime exposure to HBV infection was noted among HCW with one vaccine dose, those with no known exposure and highest in those who had Knowledge on HBsAg transmission while significant HBV protection was seen in HCW who had adequate PPE and those using gloves and dust coat consistently. A study in Southern Ethiopia found HBV lifetime exposure was higher in MWH older than 40 years(2) while population based Azar cohort study found that all age groups were exposed to HBV. In in Eastern Ethiopia, there was higher prevalence of HBV infection in trainees. (23, 24). There is need to have remedial measures that is aimed at reducing this high lifetime exposure rates by capacity building HCW, availing proper infrastructure for infection prevention and control, strengthening HBV vaccination and proper surveillance for HAI in all healthcare settings. Policy should be revised to enforce mandatory HAI pre-employment screening and vaccination for personnel working in healthcare settings.

Limitations for this study were that this was done during COVID 19 outbreak period when there was a lot of fucus on infection prevention to mitigate COVID 19 infections, in as much as this may have influenced the findings on the compliance with standard precautions, it strengthened the adherence to standard infection prevention measures. Also, during this COVID 19 period the government issued recommendations that elderly populations and those with comorbidities should work from home so we may have missed some eligible health care workers at the facilities. The study was conducted in nine highest volume government hospitals, these hospitals have the highest workload, produce the largest volume wastes, and have highest number of health care workers. We did not focus on lower level, private and faith-based health facilities where the situation could be worse or better. We only assessed HBV exposures that are related to health care settings therefore the generalizability is limited to health facilitiyrelated exposures. Health care workers are not done for thorough pre-employment medical examination so it’s difficult to point if the infections observed in this study occurred before or after employment.

## Conclusion

The prevalence of HBV infections among HCW is about 6 and 5-fold higher than general population and adolescent blood donners respectively, this high prevalence needs multi stakeholder approach to address.There was suboptimal training on waste management, infectious agent on waste and PPE coupled with PPE inadequacy that could lead to high needle stick injury poor adherence to universal/standard precautions.

There is need to ensure that adequate PPEs is available for HCW usage, trainings done to health care workers on infection prevention and control.

From the study there was low uptake of HBV vaccination with HCW immunised either due to vaccination or infection at 36.7 %, data also showed that there is significant relationship between immunization status and positivity for HBV, the high lifetime exposure may be due to high exposure to infections and low vaccination rates for personnel working in healthcare settings. Policy should be revised to make it mandatory for pre-employment HBV vaccination for most at risk populations like HCW and MWH.

No significant association was observed between HBV exposure and factors such as a history of exposure, blood transfusion, use of PPE, Knowledge on HBsAg (transmission, pathogenicity, treatment, prevention & control), training on (PPE, infectious agent, waste management). None of the sociodemographic characteristics plus other factors such as carder and departments were significantly associated with HBV exposure status for health care workers.. There is need for proper surveillance for HAI in all healthcare settings.

## Data Availability

All data collected have been reported in this manuscript

## Recommendation

There should be continuous training of HCW on universal precaution of infection prevention measures. There is need to increase HBV vaccine coverage and improve HBV surveillance among HCW.

## Acknowledgements

We acknowledge the county Government of Kisumu, each hospital’s medical superintendent and management for allowing us to do this study. We are also grateful to Kennya medical research institute, Center for Global HIV research, Human immunodeficiency Virus Research Laboratory (KEMRI,CGHR, HIV-R Laboratory) for performing the laboratory tests for the study.

We sincerely thank the data collectors and laboratory Scientists of each hospital for their assistance in data and sample collection.

We thank and appreciate the study subjects who volunteered to participate in this study. Lastly, we wish to appreciate and thank of Jaramogi Oginga Odinga University of Science And Technology for their mentor and support in designing and conducting the study.

## References

1. WHO. Global Hepatitis Report 2017_Geneva_World Health Organization_2017..pdf. 2017.

2. Amsalu A, Worku M, Tadesse E, Shimelis T. The exposure rate to hepatitis B and C viruses among medical waste handlers in three government hospitals, southern Ethiopia. Epidemiol Health. 2016;38:e2016001.

3. Anagaw B, Shiferaw Y, Anagaw B, Belyhun Y, Erku W, Biadgelegn F, et al. Seroprevalence of hepatitis B and C viruses among medical waste handlers at Gondar town Health institutions, Northwest Ethiopia. BMC research notes. 2012;5:55.

4. Franka E, El-Zoka AH, Hussein AH, Elbakosh MM, Arafa AK, Ghenghesh KS. Hepatitis B virus and hepatitis C virus in medical waste handlers in Tripoli, Libya. J Hosp Infect. 2009;72(3):258–61.

5. Atlaw D, Sahiledengle B, Tariku Z. Hepatitis B and C virus infection among healthcare workers in Africa: a systematic review and meta-analysis. Environ Health Prev Med. 2021;26(1):61.

6. Kisangau EN, Awour A, Juma B, Odhiambo D, Muasya T, Kiio SN, et al. Prevalence of hepatitis B virus infection and uptake of hepatitis B vaccine among healthcare workers, Makueni County, Kenya 2017. Journal of public health. 2018.

7. W.H.O. Progress report on HIV, viral hepatitis and sexually transmitted infections. Accountability for the global health sector strategies, 2016–2021. Geneva, World Health Organiation; 2019. Contract No.: CC BY-NC-SA 3.0 IGO.

8. WHO. Progress report on HIV, viral hepatitis and sexually transmitted infections 2019. 2019.

9. WHO. Global Health sector strategy on Viral Hepatitis 2016-2021: Towards Ending Viral Hepatitis. 2016.

10. Lok AS, McMahon BJ. Chronic hepatitis B. Hepatology. 2007;45(2):507–39.

11. Reynolds K, Thomas M, Dougan M. Diagnosis and Management of Hepatitis in Patients on Checkpoint Blockade. Oncologist. 2018;23(9):991–7.

12. Chartier Y, Emmanuel J, Pieper U, Prüss A, Rushbrook P, Stringer R, et al. WHO Library Cataloguing-in-Publication Data_Safe management of wastes from health_care activities _edited by Y. Chartier et al. _2nd ed..pdf. 2014.

13. Allegranzi B, Bagheri Nejad S, Combescure C, Graafmans W, Attar H, Donaldson L, et al. Burden of endemic health-care-associated infection in developing countries: systematic review and meta-analysis. Lancet. 2011;377(9761):228–41.

14. Shiferaw Y, Abebe T, Mihret A. Hepatitis B virus infection among medical aste handlers in Addis Ababa, Ethiopia. BMC research notes. 2011;4:479.

15. Ly KN, Kim AA, Umuro M, Drobenuic J, Williamson JM, Montgomery JM, et al. Prevalence of Hepatitis B Virus Infection in Kenya, 2007. Am J Trop Med Hyg. 2016;95(2):348–53.

16. Kisangau EN, Awour A, Juma B, Odhiambo D, Muasya T, Kiio SN, et al. Prevalence of hepatitis B virus infection and uptake of hepatitis B vaccine among healthcare workers, Makueni County, Kenya 2017. J Public Health (Oxf). 2019;41(4):765–71.

17. Otedo AE. HBV, HIV co-infection at Kisumu District Hospital, Kenya. East Afr Med J. 2004;81(12):626–30.

18. Awili HO, Gitao GC, Muchemi GM. Seroprevalence and Risk Factors for Hepatitis B Virus Infection in Adolescent Blood Donors within Selected Counties of Western Kenya. Biomed Res Int. 2020;2020:8578172.

19. Zhu S, Kahsay KM, Gui L. Knowledge, Attitudes and Practices related to standard precautions among nurses: A comparative study. J Clin Nurs. 2019;28(19-20):3538–46.

20. Sapkota B, Gupta GK, Mainali D. Impact of intervention on healthcare waste management practices in a tertiary care governmental hospital of Nepal. BMC Public Health. 2014;14:1005.

21. Aoyagi T. [ISO 15189 medical laboratory accreditation]. Rinsho Byori. 2004;52(10):860–5.

22. Mast EE, Margolis HS, Fiore AE, Brink EW, Goldstein ST, Wang SA, et al. A comprehensive immunization strategy to eliminate transmission of hepatitis B virus infection in the United States: recommendations of the Advisory Committee on Immunization Practices (ACIP) part 1: immunization of infants, children, and adolescents. MMWR Recomm Rep. 2005;54(RR-16):1–31.

23. Tesfa T, Hawulte B, Tolera A, Abate D. Hepatitis B virus infection and associated risk factors among medical students in eastern Ethiopia. PLoS One. 2021;16(2):e0247267.

24. Pouri AA, Ghojazadeh M, Shirmohammadi M, Eftekhar-Sadat AT, Somi MH. Seroepidemiology and Risk Factors of Hepatitis B Virus Infection: A Population-Based Azar Cohort Study. Iran J Public Health. 2020;49(11):2152–60.

